# Sustainable targeted interventions to mitigate the COVID-19 pandemic: A big data-driven modeling study in Hong Kong

**DOI:** 10.1101/2021.01.29.21250786

**Authors:** Hanchu Zhou, Qingpeng Zhang, Zhidong Cao, Helai Huang, Daniel Dajun Zeng

**Affiliations:** school of Data Science, City University of Hong Kong, Hong Kong, China; School of Traffic and Transportation Engineering, Central South University (CSU), Changsha, China; state Key Laboratory of Management and Control for Complex Systems, Institute of Automation, Chinese Academy of Sciences, Beijing 100091, China

**Keywords:** Agent-based modeling, Health information management, Local activities, Multilevel systems, Complex networks

## Abstract

Nonpharmaceutical interventions (NPIs) for contact suppression have been widely used worldwide, which impose harmful burdens on the population and the local economy. The evaluation of alternative NPIs is needed to confront the pandemic with less disruption. By harnessing human mobility data, we develop an agent-based model that can evaluate the efficacies of NPIs with individualized mobility simulations. Based on the model, we propose data-driven targeted interventions to mitigate the COVID-19 pandemic in Hong Kong without city-wide NPIs. We develop a data-driven agent-based model for 7.55 million Hong Kong residents to evaluate the efficacies of various NPIs in the first 80 days of the initial outbreak. The entire territory of Hong Kong is split into 4,905 500m × 500m grids. The model can simulate detailed agent interactions based on the demographics data, public facilities and functional buildings, transportation systems, and travel patterns. The general daily human mobility patterns are adopted from Google’s Community Mobility Report. The scenario without any NPIs is set as the baseline. By simulating the epidemic progression and human movement at the individual level, we proposed model-driven targeted interventions, which focus on the surgical testing and quarantine of only a small portion of regions instead of enforcing NPIs in the whole city. The efficacious of common NPIs and the proposed targeted interventions are evaluated by extensive 100 simulations. The proposed model can inform targeted interventions, which are able to effectively contain the COVID-19 outbreak with much lower disruption of the city. It represents a promising approach to sustainable NPIs to help us revive the economy of the city and the world.

## 1. Introduction

The coronavirus disease 2019 (COVID-19) pandemic has infected 116,521,281 patients and claimed 2,589,548 lives as of 6:02 pm CET, 8 March 2021 (latest information at: https://covid19.who.int/table). Various nonpharmaceutical interventions (NPIs) were implemented in response to this public health crisis worldwide, ranging from case isolation and quarantine of contacts to the complete lockdown of the entire country [1]. These NPIs were demonstrated to be able to mitigate community transmission at the expense of economic disruption, harm to social and mental well-being, and costly administration required to ensure compliance [2]. However, a resurgence of COVID-19 has been seen in many parts of the world when the NPIs were lifted [3]. As of 8 March 2021, a total of 349,398,520 vaccine doses have been administered, only account for around 4.5% of the global population. The population-wide vaccination program is not likely to cover a large-scale population in developing countries considering the huge demand and difficulty in mass production and supply chains [4], [5]. It is therefore much-needed to find the NPIs to contain outbreaks while minimizing the economic costs [6].

To provide a reliable quantitative analysis of the efficacious of alternative NPIs, we explore a data-driven targeted intervention policy to precisely identify the sub-communities that need to be surgically tested and quarantine. In this study, we develop a data-driven mobility model **(D2M2)** to characterize the demographics, public facilities and functional buildings, transportation system, and travel patterns of the 7.55 million residents in Hong Kong, an international trade center and one of the most densely populated cities in the world. Given the mass amount and rich diversity of the data publicized by the authority and public sectors, Hong Kong represents an ideal opportunity to investigate the feasibility of using a detailed agent-based severe acute respiratory syndrome coronavirus 2 (SARS-CoV-2) transmission model to inform targeted interventions. Targeted interventions informed by the model can prioritize a set of high-risk grids for quarantine, so that the localized outbreak can be contained while preserving the normal state in the majority of the city. We examine the efficacy of the proposed targeted interventions in containing the outbreak of COVID-19 in Hong Kong.

Hong Kong was one of the first places outside Mainland China to report the COVID-19 confirmed cases owing to their strong connection [7]. From January to early March 2020, the COVID-19 infected individuals in Hong Kong mainly consisted of imported cases and secondary transmissions, which is considered the first wave of COVID-19 outbreak in Hong Kong. The number of infections in the first wave remained relatively low due to a series of rapid responses, including border control, manual contact tracing, voluntary community-wide use of personal protective equipment (PPE), and social distancing [8], [9]. The second wave mainly consisted of the imported cases outside Asia and associated localized transmission, which occurred from mid-March to May 2020. After implementing more stringent NPIs, only a few local cases were reported in May and early June. On June 18, most social distancing restrictions were lifted. However, the third wave occurred in late June 2020 that pushed Hong Kong’s coronavirus case tally to 5,009 by September 19, 2020 [10]. The locally acquired cases appeared on 5 July, which is phylogenetically related to SARS-CoV-2 reported overseas [11]. There could be fourth and more waves to come, as humankind will likely to live with it until a population-wide vaccination program takes place [12]. This study is mainly to simulate the third wave outbreak in Hong Kong from June 25, 2020 to September 24, 2020.

Recently, the Hong Kong government has implemented a compulsory testing policy in response to the epidemic development and the need for infection control [13]. the government will delineate restricted areas and the people within the areas (including the person who had been present in the restricted area for more than two hours in the past 14 days) are required to stay in their premises and undergo compulsory testing and can only leave after the relevant test results are mostly ascertained. The government claims that compulsory testing policy can help to slow down the transmission of SARS-CoV-2 by early identification, early isolation, and early treatment. However, the restricted areas are identified by some index such as the occurrences of new confirmed cases or sewage samples detected COVID-19. This reactive strategy may result in a sudden directive for compulsory testing and suspension of operation of a workplace. The D2M2 can be used to forecast the epidemic dynamic and the targeted restricted areas, which could help to design a more proactive policy and inform the involved residents in advance.

The contribution of this study is threefold. First, the proposed D2M2 is calibrated by the multi-level and multi-source data of the demographics, transportation, and human mobility. To the best of our knowledge, it is the most detailed characterization of the human mobility of Hong Kong. Second, Mobile phone data is difficult to obtain in most countries. Thus D2M2, which is calibrated by open-source data, can be easily extended to the modeling of other metropolises with various demographic and human mobility patterns. Third, based on D2M2, we propose the targeted intervention, which can contain the outbreak with minimal disruption of the society. This is of particular importance to cities like Hong Kong, whose economy relies on international trade.

## II Method

### A. Synthetic Population and Their Movement Behavior

To quantitatively characterize the mobility patterns of the residents in Hong Kong, the proposed D2M2 is based on a recent human movement behavior model [14] and is calibrated with the real-world socio-demographic data of Hong Kong [15]. A human-to-human contact and transmission network can be created by combining the human behavior pattern and a synthetic population (as shown in Figure 1). Based on the demographic data from Hong Kong Population Projections and the latest Hong Kong Population Census [16], D2M2 generates synthetic agents with the attributes of age, occupation, ethnicity, and gender. Hence, a total of 7,550,000 agents are generated for Hong Kong residents, of which 5.14 million are individuals between 20 and 65, 9.8 million are students, and the rest are other agents such as elder people and infants. Then each agent is allocated to a corresponding family according to the pre-specific family structure for 2.66 million households. Households were geolocated within 214 discrete areas named Tertiary Planning Units (TPUs) according to spatial characteristics outlined in the census and the TPUs are further split into 4,905 500m × 500m grids for better aggregated the spatial characteristics of households.

**Fig. 1.**
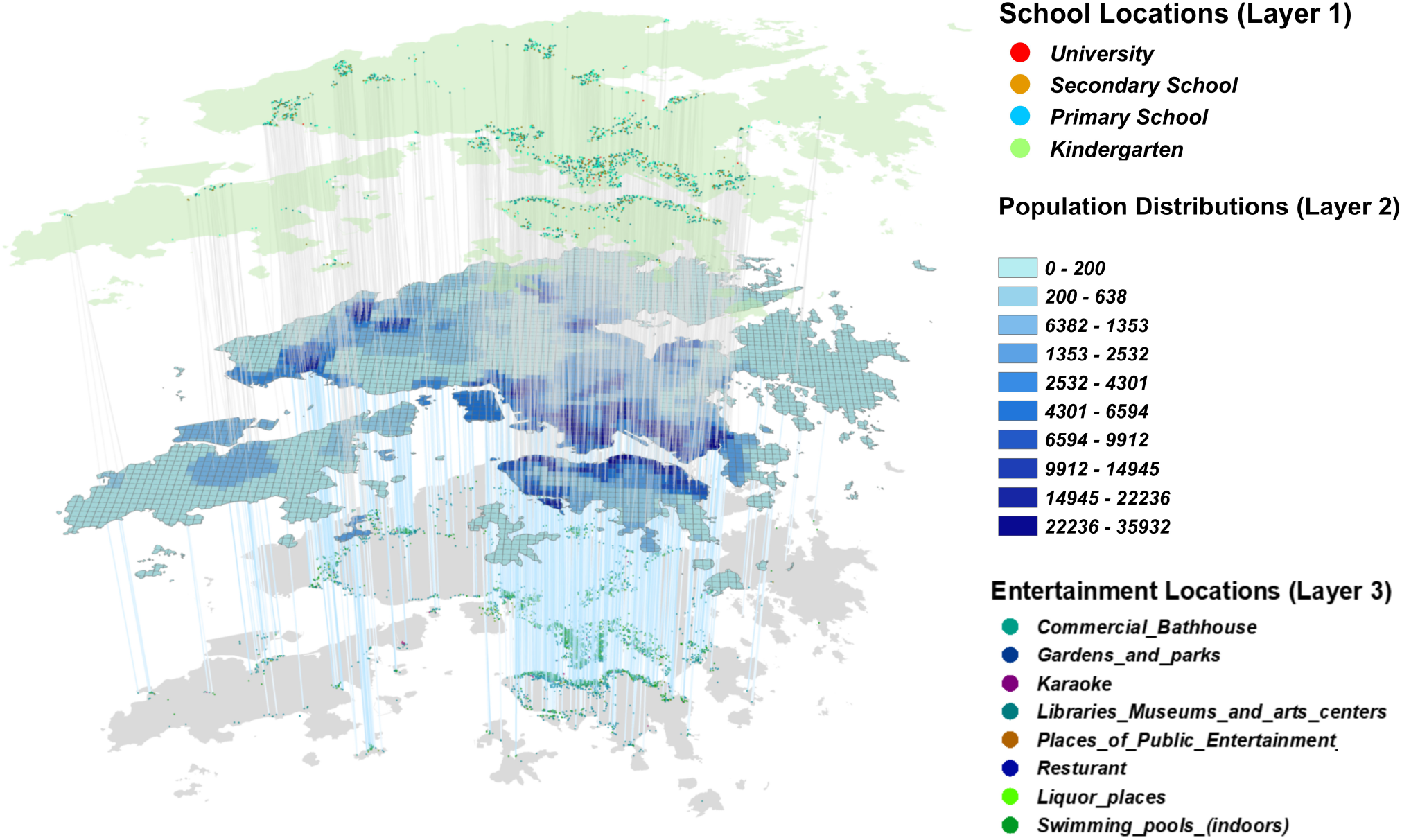
Geographical distribution of the synthetic population and facilities in Hong Kong. The D2M2 comprises 5.14m adults and 0.92m students and 1.49m other agents (infants or elderly people).

The agents interact in a comprehensive set of Points of Interest (POIs), including houses, workplaces, schools, restaurants, leisure and cultural services venues, shopping malls, pharmacies, groceries, various public entertainment places, etc. The agents interacted at home and community, workplace, school, and other functional buildings. According to the setting previous study [14], during the night, we assume all agents back home and interacted primarily within their households regardless of the weekday or weekends. During the daytime weekday, the agents were divided into three groups: the commuters (i.e., the people who are not working at home) interacted with individuals in the same workplace, and students within their schools while the home-staying agents interact with the individuals who live in the same house or community. In the daytime of weekends, all agents interacted with others according to the spatial distribution of different recreational places (as shown in Figure 2). In addition, the Foreign Domestic Helpers (FDHs) represent an integral part of Hong Kong society nowadays, and 5.29% of the population [17]. We set 39,9320 FDHs among the home working agents according to the statistics of the Immigration Department in 2019 [18]. We set FDHs gather on Sundays in public areas [19], and thus represent the potential risk of localized outbreaks.

**Fig. 2.**
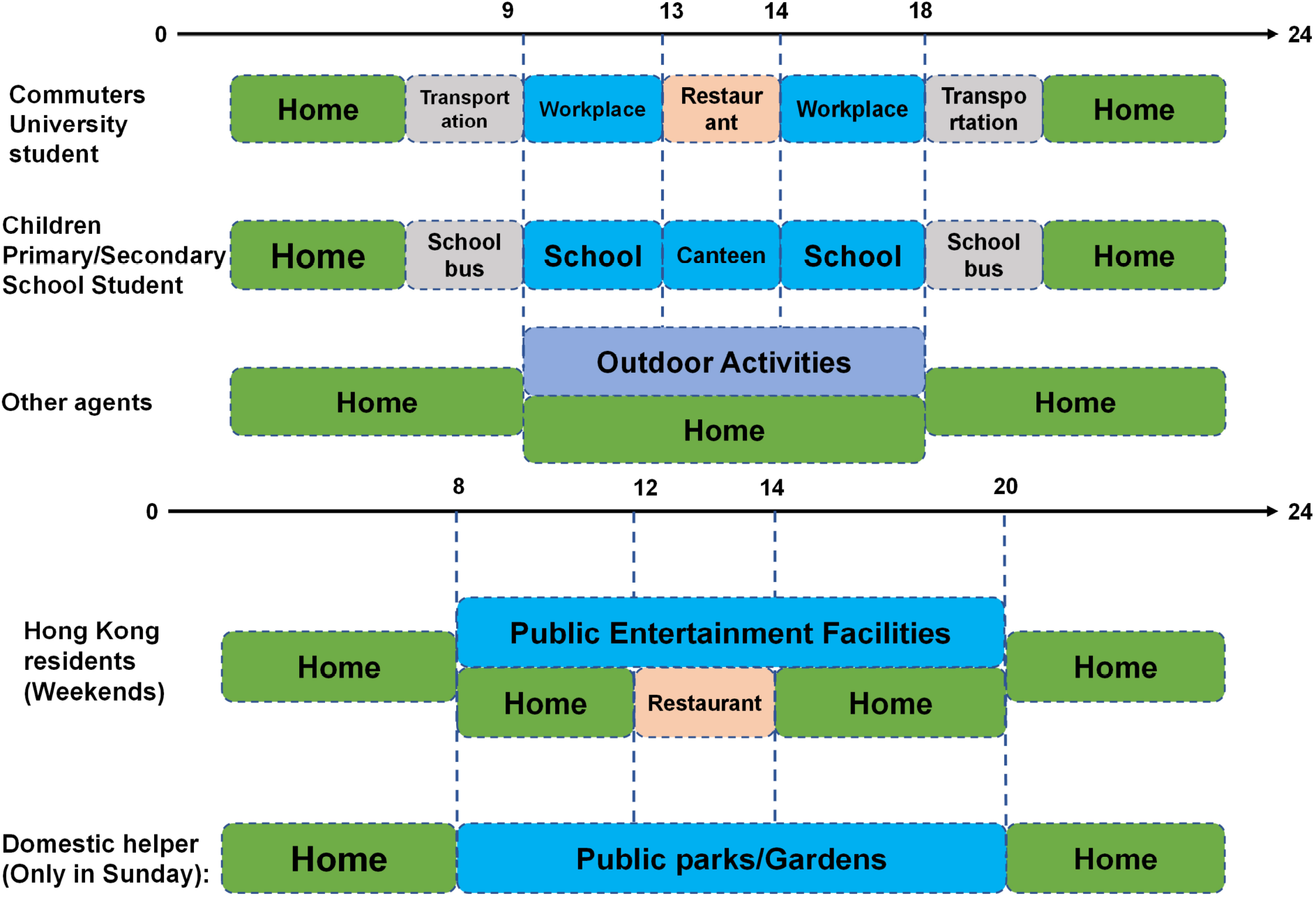
Schematic illustration of the daily movement of three groups of people in Hong Kong on weekdays and weekends. Commuters include people aged 23–60 who work outside home or study at universities. Students includes children and adolescents aged 4–18 who are studying at schools (excluding university); Others includes children less than 4 years of age, retired elderly adults **(> 65)**, the unemployed, and those who work from home.

### B. Transmissiom Model

In the transmission model, infection likelihood between agents is related to the time that he or she spent where infected agents have visited [20].Suppose *i* and *j* are two agents. *j* is infectious, we denote the probability of *j* infecting *i* on day *t* in location type *l* as:

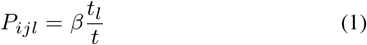

Here *β* is the likelihood for individuals to infect one another at specific places and the wider TPU, which is obtained from the basic reproduction number [2], [20]. *t*_*l*_ is the duration of exposure to infection, *t* is a constant for indoor hours per day. Hence, the probability of individual *i* getting infected from the location *l* on day *t* can be expressed as:

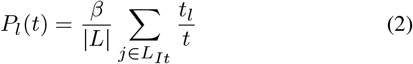

where *L* is a set of individuals of location *l, L*_*It*_ is the subset of all individuals who belong to set *L* and are infectious on day *t*. |·| denotes the size of the set of individuals and |*L*| represents the total number of agents in set *L*. The total number of agents infected in location *l* on day *t X*_*L*_(*t*) can be denoted by a random process:

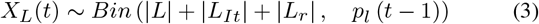

where *L*_*r*_ is the subset of *L* consisting of agents that have been removed as a result of death or recovery.

### C. Model Parameterization and Fitting

A recently introduced susceptible–latent–infected–removed (SLIR) model with some additional compartments is adapted to incorporate the unique characteristics of COVID-19 (as shown in Figure 3) [6], [20], [21]. In particular, at each time step *t* (one day), the infectious asymptomatic (*I*_*A*_), infectious symptomatic (*I*_*S*_) and pre-symptomatic (*P*_*S*_) individuals can transmit thevirus to susceptible (*S*) individuals with probability *rβ, β* and *β*_*s*_, respectively. If the transmission is successful, the susceptible agent will move to the latent asymptomatic state with probability *p* or to the latent symptomatic state (*L*_*S*_) with probability (1−*p*). A latent asymptomatic individual becomes infectious asymptomatic after a period *ϵ*^′−1^, and some latent symptomatic individuals will transit to pre-symptomatic (*P*_*S*_) state after the same period. Then, during *μ*^−1^ period after being infected, the symptomatic individuals become onset cases and can either recover or removed after a period *μ*^−1^.

**Fig. 3.**
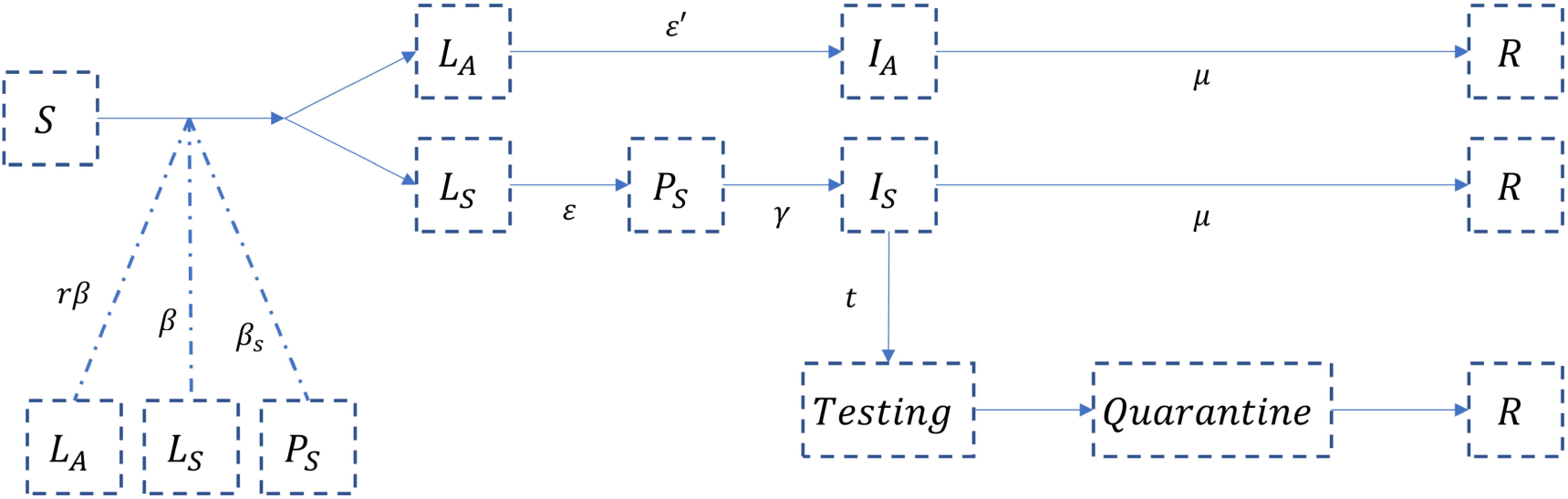
The SLIR model. Here we denote susceptible **(*S*)**, latent asymptomatic **(*L***_***A***_), latent symptomatic **(*L***_***S***_), presymptomatic **(*P***_***S***_), infectious asymptomatic **(*I***_***A***_), infectious symptomatic **(*I***_***S***_), testing symptomatic case **(*T esting*)**, positive in testing and quarantine **(*Quarantine*)** and recovered/removed **(*R*)** individuals.

The values for the basic reproduction number (*R*_0_) were set as 2.5 according to the literature [22], [23]. Following the setting of the previous study, we ran the models for 80 days to investigate the early stages of a localized outbreak and randomly selected 50 agents as the initial infected on day 0 [20]. The parameters used in the model from literatures are presented in Table I.

**TABLE I.** The parameters used in the transmission model

| Parameters | Description | Value | References |
| --- | --- | --- | --- |
| $\gamma$ | Relative infectiousness of asymptomatic individuals | 50% | [24] |
| $\epsilon^{-1}$ | Incubation period | 6.2 days | [25] |
| $\epsilon'^{-1}$ | Latent period (for asymptomatic and symptomatic case) | 2.9 days | [21] |
| $pre$ | pre-symptomatic period | $\epsilon^{-1} - \epsilon'^{-1}$ days | [21] |
| $p$ | Proportion of asymptomatic | 17.7% | [26] |
| $\mu^{-1}$ | Time to removed/recovery | 2.9 days | [23] |
| $k$ | Proportion of presymptomatic transmission | 12.6% | [27] |
| $R_0$ | Basic reproduction number | 2.5 | [6] |
| $\beta$ | Basic transmission rate | $\frac{R_0 \mu}{pr + (1-p) \div (1-k)}$ | [6] |

To validate the utility of the model set, we simulate the third wave of COVID-19 epidemic in Hong Kong, which began on 25 June 2020 and has been contained in late September after a hard-fought battle. We set the starting date of the simulation to be 25 June 2020 when a cook in a local restaurant had the onset of symptoms, breaking the 21-day streak without local infections. We set the ending date to be 25 September 2020 as the Chief Executive of Hong Kong S.A.R. Government said the third wave of the COVID-19 epidemic has been contained. The infection probability at different locations is being updated based on the real-time responses of the government. After the Education Bureau announced *school closure* on 13 July [28], we assume the infection probability at school to be 0 and the residents’ infection probability at other locations are being updated according to Google’s Community Mobility Reports. On 19 July, the Hong Kong S.A.R. Government announced a series of anti-pandemic measures such as the bans on dine-in services at restaurants from 6 pm to 5 am (*anti-pandemic measures*). On 27 July, the government further tightened social distancing measures, including gathering in public will be limited to only two people per group and restaurants are disallowed to offer dine-in services (*tightened measures*). These measures may result in a huge decrease in contact intensity. Thus, based on the previous studies and surveys [29], we assumed that on 19 July, the contact intensity decreased to 50% and then further decrease to 10% on 27 July of the normal situation at the workplace and other public places respectively. Moreover, we also assumed the people who live or work with someone diagnosed with COVID-19 will follow the Home Quarantine Rule [30]. These agents will complete the 14 full days of self-isolation (separate themselves from others at home) and cannot infect other agents.

### D. Targeted Interventions, Social-distancing, and Testing Strategies

To confront the pandemic with less disruption, we propose the *targeted interventions*, which identify a small subset of areas for quarantine while maintaining mobility in other areas. Specifically, we run 200 simulations of *baseline scenario* (i.e., no NPIs) with 50 randomly seeded agents. Then,following Koo’s study [20], based on the outcome of the baseline scenario simulations at day 80, thegrids are ranked by the number of expected infections in the baseline simulation. The top 50 and top 100 grids are chosen as the High-Risk Grids (HRGs), while the others are non-HRGs. Compulsory quarantine is taking place at HRGs. Within HRGs, all essential materials are uniformly delivered by the authorities. Residents in non-HRGs are not affected except that they could not freely commute to HRGs. As a comparison, we examine the efficacy of the *stay-at-home control*, which involves the following scenario starting from day 15 (the actual delay in the response in Hong Kong [31]): (a) School closures. We transform all school students to home working agents simultaneously. (b) Social distancing. We assume that all places are still open, but the volume of passengers is proportional to the Google Mobility Report on the same day.

Moreover, we examine the efficacy of the *reactive control*. Testing is a useful tool to detect, predict, and reduce the spread of disease [32], [33]. In this scenario, in addition to the *stay-at-home* control, patients with symptoms are quickly tested and quarantined since the authorities respond on day 15 [34]. We set that testing results will come out either 24 or 48 after being tested [6]. In the experiment, we evaluate the outcomes with different proportions of symptomatic patients who get tested(10%, 20%, 50%). The maximum testing capacity is set as 10,000 cases per day according to the governments’ publication [35]. Finally, we investigate the *full lockdown control* on the top of testing 50% of symptomatic cases, which represents the scenario that school closer the 70% reduction in human mobility is taken place on day 1, which follows the setting of strict lockdown in the study of US National Bureau of Economic Research [36]. Note that we run 100 simulations for non-baseline NPIs (summarized in Table II) to account for the stochasticity and to calculate the confidence intervals (CIs).

**TABLE II.** Summary of applied NPIS

| NPIS | Description |
| --- | --- |
| *Baseline* | No intervention |
| *Stay-at-home control* | School closure + Mobility changed |
| *Reactive control* | *Stay-at-home control* + testing 10%, 20% or 50% of symptomatic patients |
| *Full lockdown control* | *Strictest reactive control* + 70% mobility reduction |
| *Targeted interventions* | Controlling 50 or 100 girds facing the highest infection risk |

## III Results

### A. Compare the Simulation with Real-world Data

Figure 4 presents the comparison of the simulation results of the proposed D2M2 model with a random number of initial infections (5 to 15) and the number of confirmed cases in Hong Kong during the third wave. Note that we use the onset date instead of the reported date of each infected case and exclude the asymptomatic cases to reflect the real situation. In general, the simulation results (blue curve) capture the trend and turning point of the confirmed case data (grey bars). The real-world data are within the interquartile range (IQR) of the model simulations. The blue curve (the median of simulations) is a little lower than the real-world data because of the superspreading events, which are likely to result in more explosive growth [37]. Specifically, our model is a theoretical model that simulates viral transmission based on the general population movement pattern and average disease transmissibility, and thus lack of data from large clusters or superspreading events to simulate the real-world situations [38]. The implementations of the control measures are represented by the red lines. The effects of anti-pandemic and further tightened measures became obvious with fluctuations, due to the incubation period and asymptomatic transmission. Moreover, the effect of school closure just slightly affects the infection number.

**Fig. 4.**
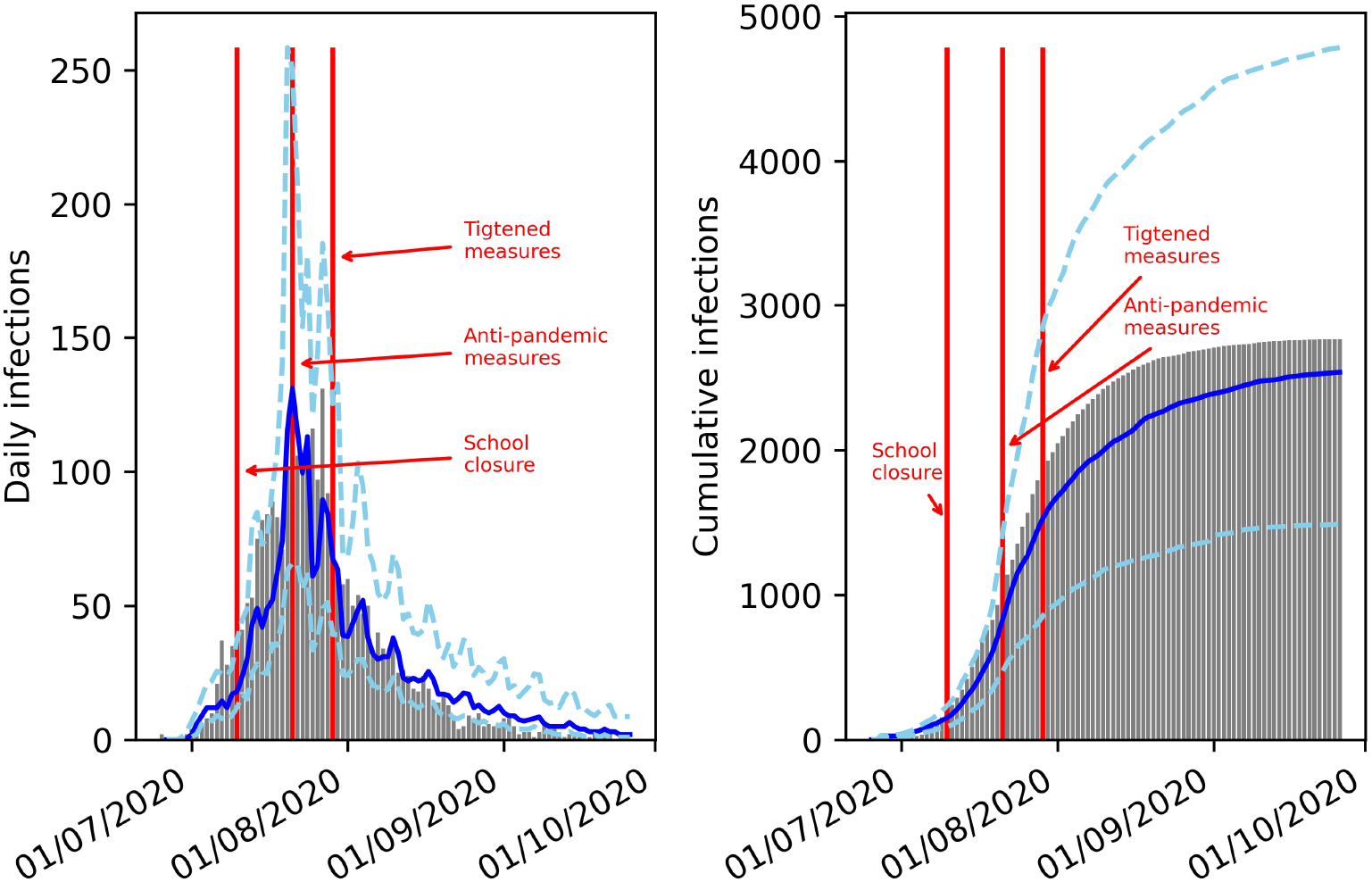
Comparison of simulated results with the official confirmed case. The total number of daily infections is shown on the left; the cumulative number of infections is shown on the right. The blue curve (dashed curve for IQR) and grey bars represent the simulated results and the confirmed cases, respectively. The red vertical lines indicate the actual control measures in Hong Kong.

### B. Quantify the Effectiveness of NPIs

For the baseline scenario, a cumulative median of around 129,000 cases (IQR: 94,000-193,000) of the Hong Kong population was infected on day 80 (Figure 3, Table III). At this level of transmission (*R*_0_=2.5), mild social distancing resulted in a median of around 17,000 cases (IQR: 12,000–25,000), the full lockdown (city-wide quarantine) measure led to 805 cases (IQR: 636–10,60). Regrading reactive control measure with a 24-hour-delay, testing 10% of onset cases led to in a median of around 13,000 cases (IQR: 9,000 - 21,000); testing 20% of onset cases led to 5,830 cases (IQR: 4,315-9,204); Testing 50% led to 2,667 cases (IQR: 2,187-3,618). With a 48-hour-delay, the infected cases number resulted by testing 10%, 20%, and 50% symptomatic cases would be 16,000 (IQR: 9,000-31,000), 9,761 (IQR: 7,584-17,675) and 5,649 (IQR: 3,691-9,433) respectively. Compared with the baseline scenario, the full lockdown led to the greatest reduction in cases (99.32%, IQR: 95.83%-99.37%).

The location of the infection (school, workplace, domestic party, non-commuter place or home community) was set as the site where individuals acquired the virus from others. As shown in Table 3, in most scenarios, the home was the place with the highest number of infections. Workplace contributed the second-highest number of infectious, indicating that reducing the mobility in the workplace could reduce the risk of infections since the commuters interacted with agents in the same workplaces or functional buildings [20]. School closure could reduce the infectious among students but contributed little to protect other residents. This has also been confirmed by the previous study [29]. Moreover, the number of infections at FDHs’ gathering of other POIs was relatively consistent and count small proportions in different NPIs. In the full lockdown scenario, the infections mainly occurred at the POIs that are neither school or workplace, including grocery, pharmacy, or retail store. This indicates we may further reduce the number of infections by reducing the activities in POIs at the cost of wider social disruption.

**TABLE III.**
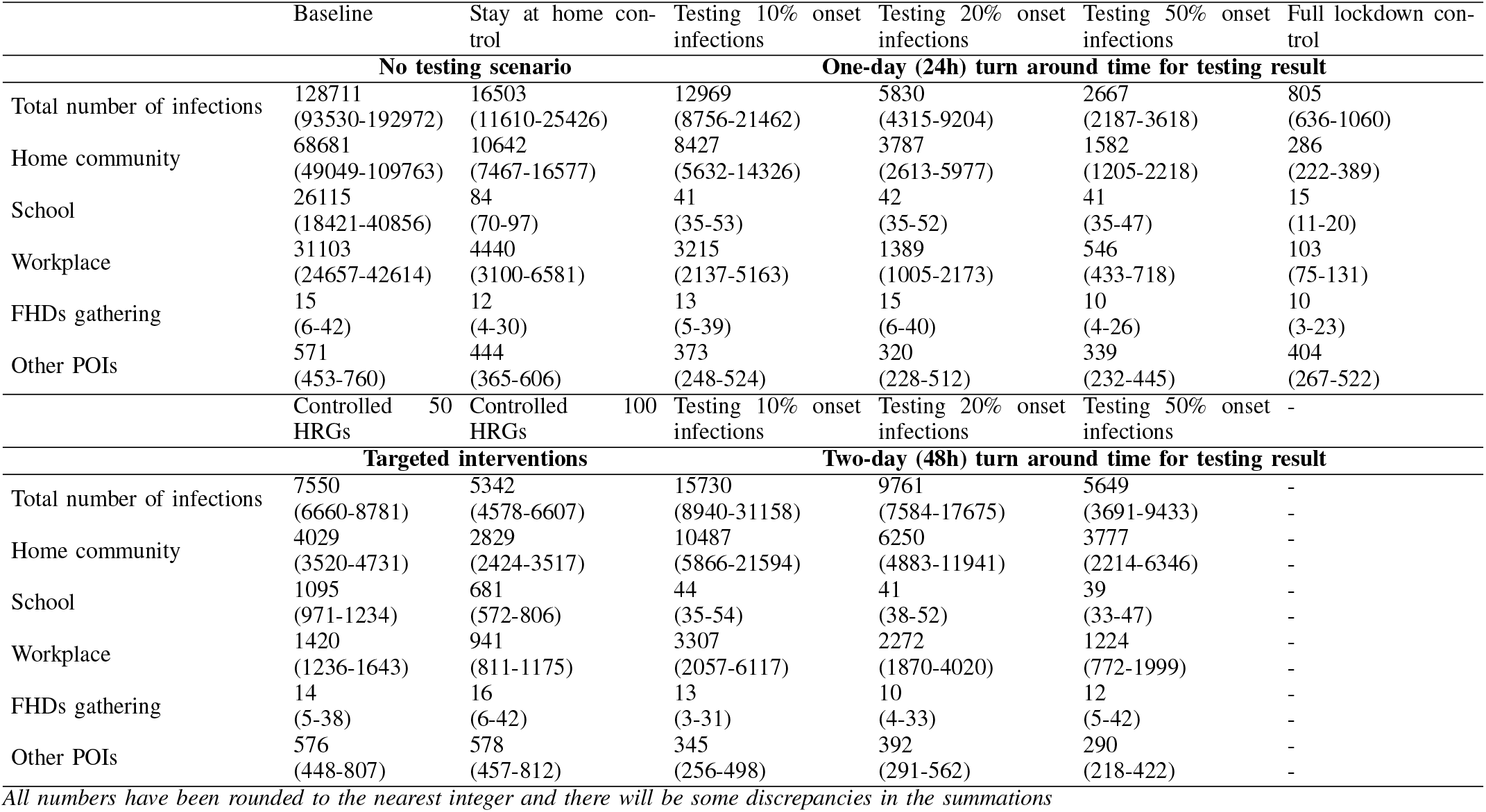
Estimated median (IQR) or cumulative number of SARS-COV-2 infections on day 80 by location, intervention, and testing results delay

The epidemic peak has not been reached by day 80 in all scenarios, because here we did not consider the contact tracing for model simplicity. As shown in Figure 3, the median number of newly infected cases in the baseline and mild social distancing scenarios was consistently increasing, whereas the number of new cases was relatively stable under the instantaneous response. From the spatial perspective, a considerable number of new infections were acquired from the urban areas in Kowloon and New Territories (Figure 5). The instantaneous response can effectively reduce the spillover effect of the epidemic outbreak from urban to suburban area. The infection distributions in the baseline and mild social distancing scenarios were more widely distributed in less population-dense areas. Given the dense residential clusters mainly sited on the Kowloon, Hong Kong island, and the new towns of New Territories, the Large-scale and homogeneously distributed infections implied that the virus cannot be contained without massive testing or total lockdown strategy.

**Fig. 5.**
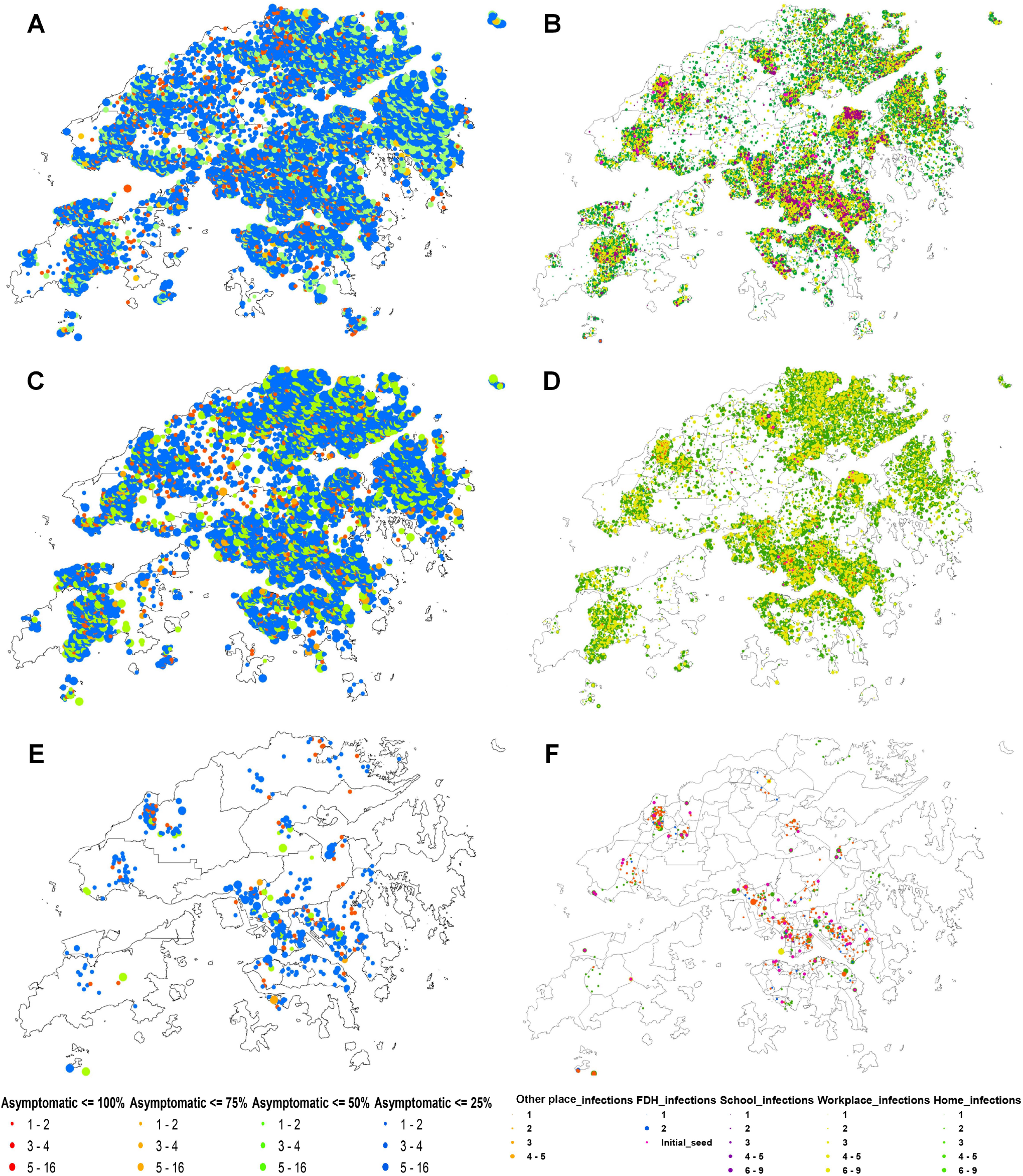
Geospatial distribution of the simulated infections on day 80. The locations are determined by the household location of infected individuals. The size of dots is proportional to the number of infections in the corresponding household. For panels A, C, and E, the color of dots represents the percentage of asymptomatic cases in each household. Blue: 0%-25%; Green: 25%-50%; Orange: 50%-75%; Red: 75%-100%. For panel B, D, and F, the color of dots represents the source of the infection. Green: household; Yellow: workplace; Purple: school; Blue: gathering of domestic helpers; Orange: Other POIs.

### C. Results of Targeted Interventions

The targeted interventions were derived as follows. First, we ran simulations of the baseline scenario (Figure 7). Second, the top 100 and 50 grids with the highest number of infections were chosen as the HRGs (Figure 8), which will be quarantined, whereas the people who live in other non-HRG grids were not affected. Therefore, the targeted interventions are proactive NPIs, which are based on the prediction of the epidemic patterns. Simulation results showed that the targeted interventions could effectively reduce the size of the outbreak in the city, with 5,342 (IQR 4,578-6,607) and 7,550 (6,660-8,781) accumulative infected individuals after 80 days by only quarantining 100 (2.04% of the area and 24.46% of the population) or 50 (1.02% of the area and 13.05% of the population) grids in the city. This indicated that the COVID-19 outbreak was largely contained without much disruption of society. The efficacy of quarantining 100 HRGs slightly outperformed the reactive control strategy with 50% symptomatic cases tested within 48 hours (Figure 9). Considering the high cost of contact tracing and mass testing, the targeted interventions stand a good potential to become reliable and cost-efficient NPIs to confront the long-term COVID-19 pandemic and even endemic.

**Fig. 6.**
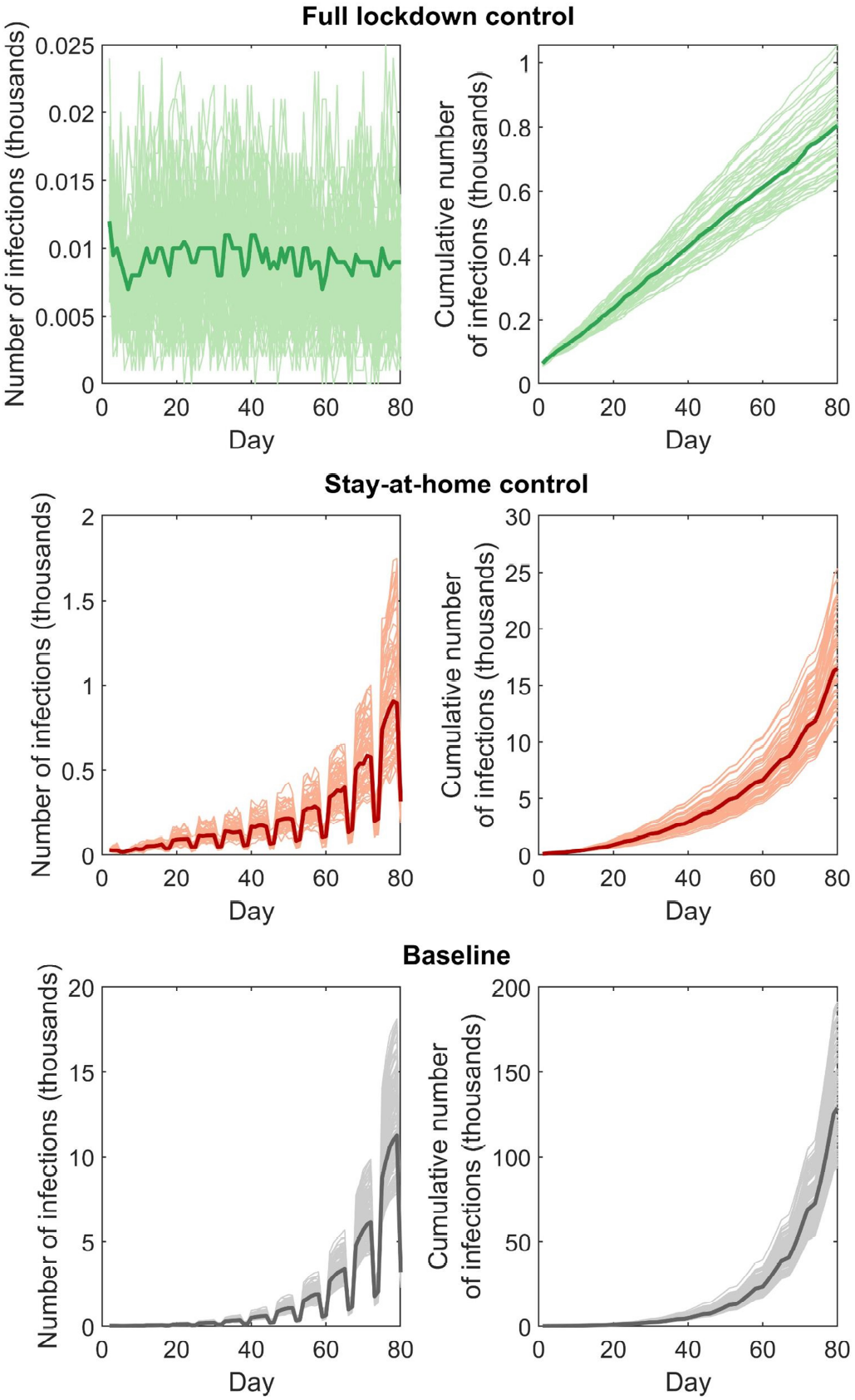
Total daily number and cumulative number of SARS-CoV-2 infections up to 80 days after different control measures Total number of daily infections is shown on the left; cumulative number of infections is shown on the right. Dark lines represent the medians in each panel. Shaded areas represent the MC simulation results.

**Fig. 7.**
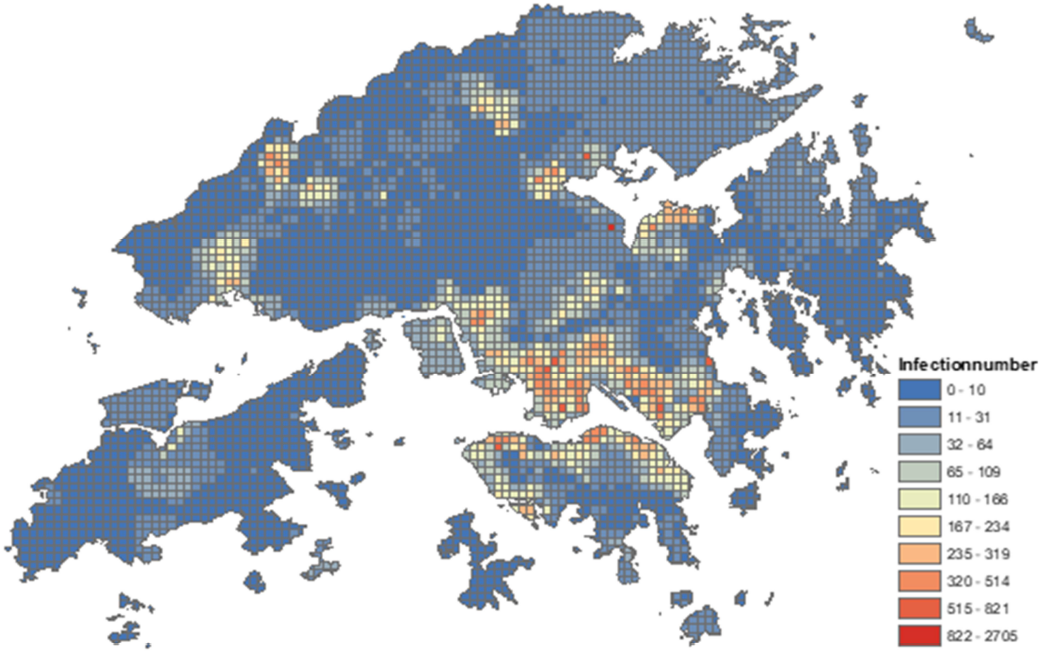
The heat map for the infections across the city in the baseline scenario (no NPIs).

**Fig. 8.**
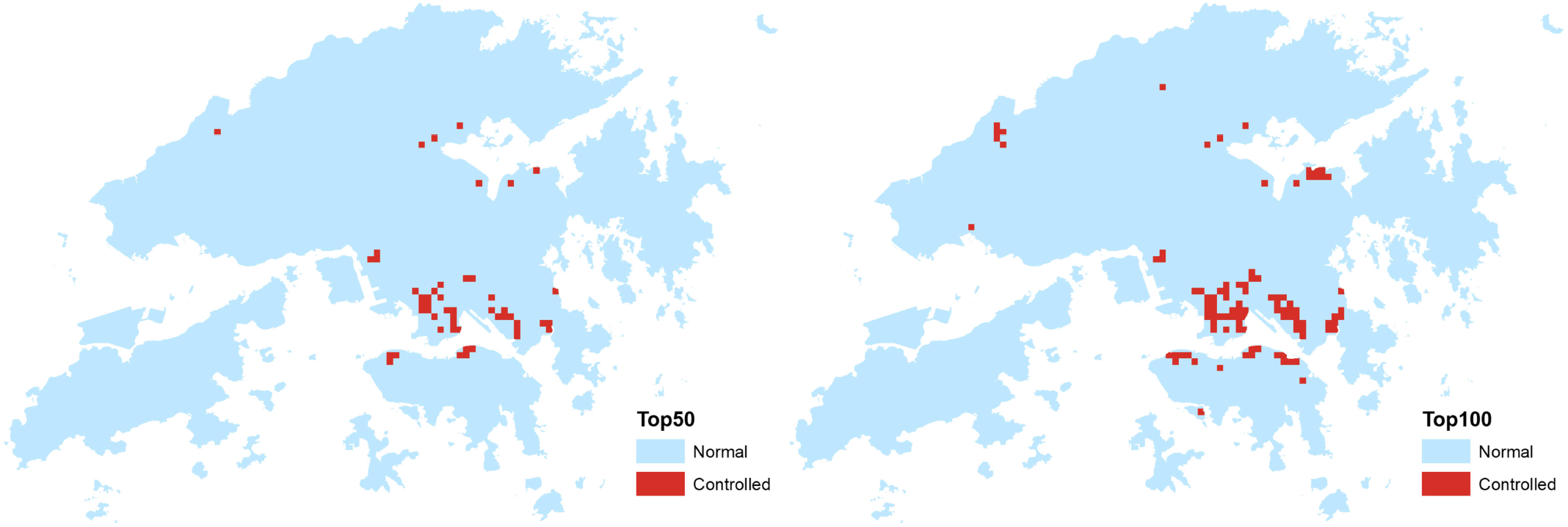
The geospatial distributions of identified HRGs (left: top 50 grids, right: top 100 grids).

**Fig. 9.**
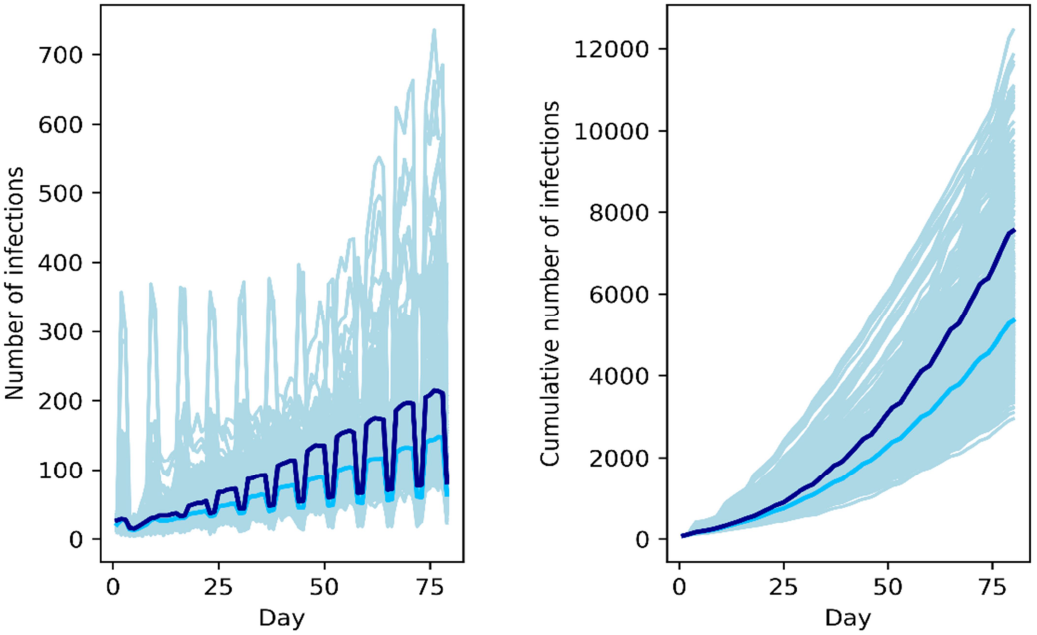
Results of the targeted interventions. The total number of daily infections is shown on the left; cumulative number of infections is shown on the right. The dark navy lines represent the corresponding median when 50 grids were controlled. Light blue lines represent the medians in each panel when 100 grids were controlled. Light blue shading represents all 200 simulations for each scenario.

**Fig. 10.**
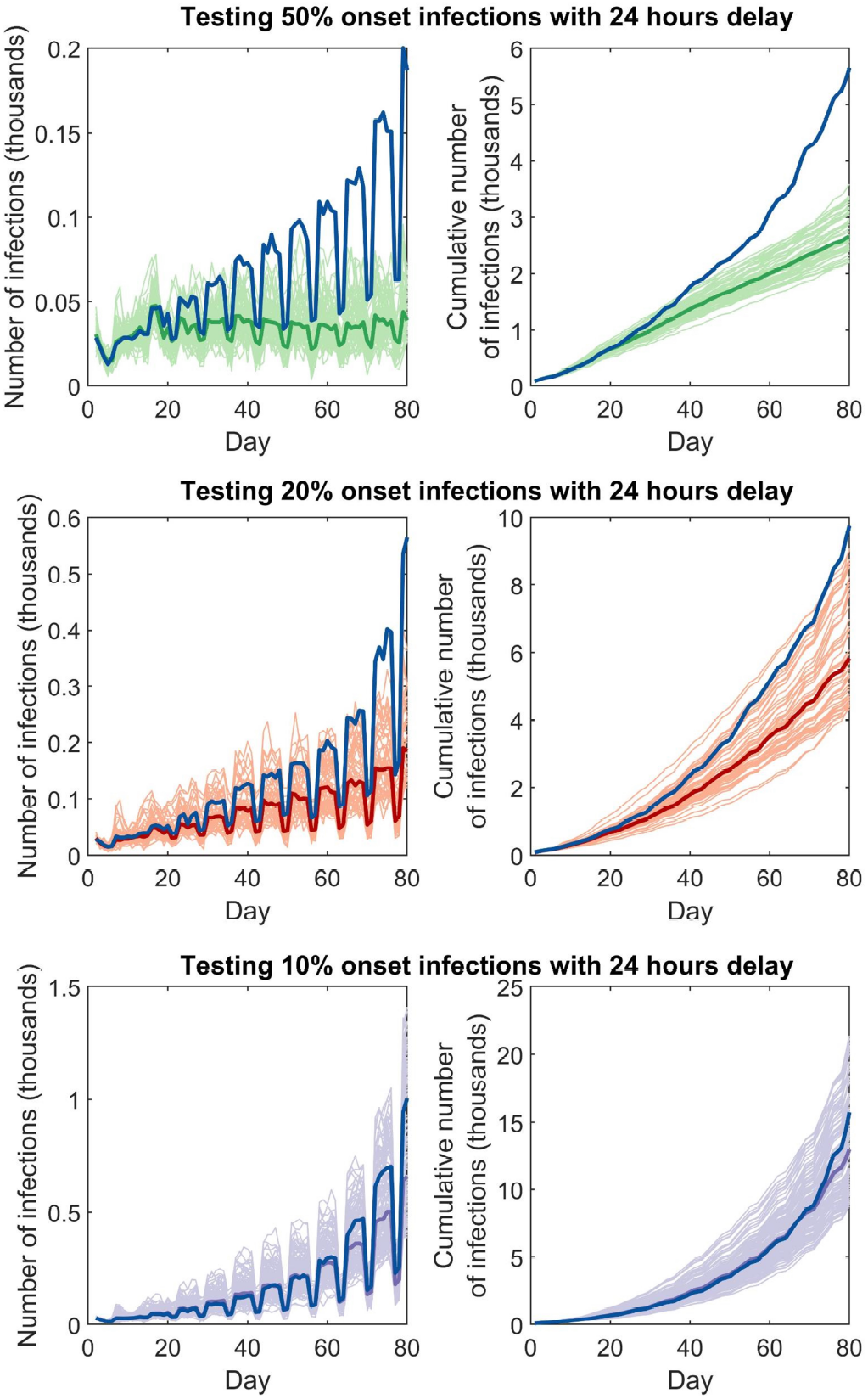
Total daily number and cumulative number of SARS-CoV-2 infections up to 80 days after different 24 hours delay testing scenarios. Total number of daily infections is shown on the left; cumulative number of infections is shown on the right. Dark green, dark orange and purple lines represent the median number when there 24 hours delay for tests. Dark navy lines represent the corresponding medians of 48 hours output delay as a comparison. Shaded areas show all 100 simulations for each scenario.

Geospatially, the top 50 HRGs are mainly located in urban areas in Kowloon and New Territories, two of the most densely populated districts in the world. More grids in Hong Kong Island were identified as in the top 50-100 HRGs. The targeted interventions had a suppressive effect on new infections over time, though the number of cases continued to increase, and the cumulative number of cases are exponentially growing, implying a delayed intervention effect rather than a preventive effect here as observed in the full lockdown scenario.

### D. Results of Reactive Control Measure

We examined the effect of testing by simulating three scenarios that 10%, 20%, and 50% percent of symptomatic cases are tested after the first 15 days (Figure. 6). Moreover, to analyze the effect of testing speed, the testing results are set to come out 24-hour or 48-hour after the testing. Compared with the baseline data, testing 10%, 20%, 50% of symptomatic cases within 24 hours reduced the infectious by 89.92% (IQR 88.88%-90.64%), 95.47% (95.23%-95.87%) and 97.93% (97.66%-98.13%), respectively. As contrast, testing 10%, 20%, 50% of symptomatic cases within 48 hours reduced in 87.78% (83.85% - 90.44%), 92.42% (91.89%-92.84%) and 95.61% (95.11% -96.05%), respectively. Apparently, the speed of testing greatly affects the efficacy of NPIs. These results indicate that timely and prompt testing is essential to such reactive control measure [39].

Regarding the location of infections, the home-community and workplace ranked 1st and 2nd in all testing scenarios. More importantly, due to the stay-at-home and testing strategies, the home-community was responsible for more than half of infections (Table III). In the scenario of 24-hour testing delay, the home-community and workplace infections accounted for 89.76% (88.73%-90.81%), 88.78% (83.84%-88.93%) and 79.79% (74.89%-81.14%) at 10%, 20% and 50% testing scale level, respectively. The corresponding values were 88.62% (88.09%-88.93%), 89.04% (87.31%-90.30%) and 88.52% (80.89%-88.66%) in the scenario of 48-hour testing delay. Compared to the full lockdown scenario, the wide gap between the infection sizes implied that the community and workplace-based transmission is dramatically growing and became hard to control if no strict movement restrictions were implemented during the outbreak period.

## IV. Discussion

COVID-19 has resulted in enormous damage to the society and economy. Many countries have been suffering from a shortage of testing resources and economic stagnation due to the necessary lockdowns. The quantitative analyses with D2M2, which is calibrated with the real-world big data of human mobility, demonstrate that targeted interventions are feasible NPIs to contain the outbreak with only a small fraction of areas and population being affected.

Apparently, the most effective strategy to contain the COVID-19 outbreak is to have a stringent lockdown, which is also echoed by our results. However, city-wide quarantine and lockdown may lead to additional loss including the elevating unemployment rate, social isolation, and mental health crisis [40], [41]. Moreover, COVID-19 has the potential to become an endemic [12], if long-term protection of vaccines is not satisfactory. Therefore, many countries would have to lift the NPIs with a certain level of COVID-19 risk for the near future [42]. In practice, the proposed targeted intervention strategy can partially mitigate isolation fatigue and social disruption by proactively identifying only a small subset of the area and population for testing and quarantine, while preserving the mobility of most residents. Furthermore, as an agent-based model, D2M2 is versatile and can be modified to incorporate other factors such as vaccination programs.

In practice, as an international financial center and a major supply chain hub, Hong Kong is likely to face the continuous risk of COVID-19 in the near future. The proposed D2M2 and associated targeted interventions have the potential to guide the current compulsory COVID-19 testing policy achieves a balance between lowering the risk and preserving the human mobility and economy of the city. This also applies to other major cities in the world, such as Beijing, New York, London, Tokyo, etc. In the big data era, it is essential and promising to capitalize on the well of valuable data about human mobility and urban structure to benefit mankind in various contexts. Our study is an effort in combining real-world data of population demographics, public facilities and functional buildings, transportation systems, and travel patterns to inform effective NPIs to combat emerging infectious diseases like COVID-19. The limitation of this study is the missing of real-time individualized mobility movement data such as the mobile phone data. Obtaining individualized mobile phone data is extremely difficult, if not impossible. However, the model can be applied to other cities where such individualized mobility data is available.

## Data Availability

Custom code that supports the findings of this study is available from the corresponding author upon request.

---

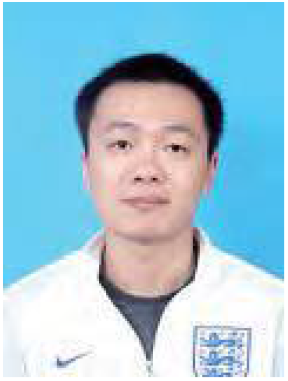

**First A. Author** received the Bachelor’s degree in Traffic Engineering and Software Engineering from Dalian Jiaotong University, Liaoning, China, in 2014. And received the master’s degree in Communication and Transportation Engineering from Central South University, Hunan, China, in 2017. He is currently working toward a joint PhD degree with Central South University, Hunan, China and City University of Hong Kong, Hong Kong, China. His research area includes Data analysis,intelligent transportation systems, and statistical analysis

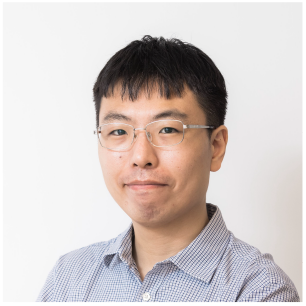

**Qingpeng Zhang** received the B.S. degree in automation from the Huazhong University of Science and Technology, Wuhan, China, in 2009, and the Ph.D. degree in systems and industrial engineering from the University of Arizona, Tucson, AZ, USA, in 2012.

He was a Post-Doctoral Research Associate with the Department of Computer Science, Tetherless World Constellation, Rensselaer Polytechnic Institute, Troy, NY, USA. He is currently an Associate Professor with the School of Data Science, City University of Hong Kong, Hong Kong. His current research interests include medical informatics and network science. Dr.Zhang is an Associate Editor of IEEE Transactions on Intelligent Transportation Systems, IEEE Transactions on Computational Social Systems, Journal of Alzheimer’s Disease, and PLoS ONE.

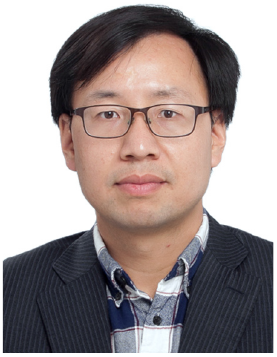

**Zhidong Cao** received the Ph.D. degree in geographic information science from the Institute of Geographic Sciences and Nature Resources Research, Chinese Academy of Sciences, Beijing, China, in 2008. He is currently an Associate Professor at the Institute of Automation, Chinese Academy of Sciences. His current research interests include social computing, public health, emergency management, and spatial analysis.

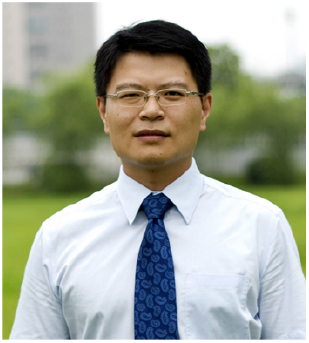

**Helai Huang** Dr. Helai Huang is a professor of transportation engineering in the School of Traffic and Transportation Engineering,Central South University (CSU). He serves as associate dean of the School, the founding director of CSU Urban Transport Research Centre and Key Laboratory of Smart Transportation of Hunan Province. Dr. Huang holds B.E and M.S. degrees from Tianjin University, China (1996-2003), a PhD degree in transportation engineering from National University of Singapore (2003-2007), and worked as a post-doc at University of Central Florida (2008-2010).

His research interests include traffic safety, transportation planning and ITS. Dr. Huang is a Chief Editor of Accident Analysis and Prevention (Elsevier), an editorial board member of Analytic Method in Accident Research (Elsevier) and Journal of Geography and Regional Planning, a committee member for TRB ABJ80 Committee (Statistical Methods) and TRB ANF10 Committee (Pedestrians), and a journal reviewer for over 20 SCI/SSCI -indexed international journals.

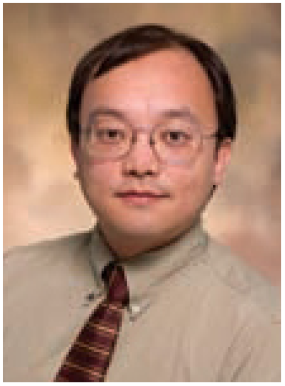

**Daniel Zeng** received the B.S. degree in economics and operations research from the University of Science and Technology of China, Hefei, China, and the M.S. and Ph.D. degrees in industrial administration from Carnegie Mellon University. He is Gentile Family Professor in the Department of Management Information Systems at the University of Arizona. He also holds a Research Fellow position at the Institute of Automation, Chinese Academy of Sciences. His research interests include intelligence and security informatics, infectious disease informatics, social computing, recommender systems, software agents, and applied operations research and game theory. He has published more than 300 peer-reviewed articles. He currently serves as the editor in chief of ACM Transactions on MIS, and President of the IEEE Intelligent Transportation Systems Society. He is a Fellow of IEEE.

